# Whole exome sequencing identifies genes associated with non-obstructive azoospermia

**DOI:** 10.1101/2020.05.29.20116558

**Authors:** Hongguo Zhang, Hui Huang, Xinyue Zhang, Wei Li, Yuting Jiang, Jia Li, Leilei Li, Jing Zhao, Jing He, Mucheng Chen, Ruixue Wang, Jing Wu, Zhiyu Peng, Ruizhi Liu

## Abstract

**Background:** Genetic etiology is the main cause of non-obstructive azoospermia, but little is known about the landscape of the disease causative genes.

**Objective:** To identify the association of non-obstructive azoospermia and the putative causative genetic factors.

**Design, setting, and participants:** A single-center perspective case-control study of 133 patients, with clinicopathologic non-obstructive azoospermia and 495 fertile men control was performed. Eleven trio families were available and enrolled from the 133 patients’ families.

**Outcome measurements and statistical analysis:** Whole exome sequencing based rare variant association study between the cases and controls was performed by means of gene burden association testing. Linkage analysis on the trio family was also described to screen the causative genes.

**Results and limitations:** Totally 80 genes (p < 0.05) were identified associated with non-obstructive azoospermia (2 of which were previously reported), meanwhile 5 novel genes out of which were also found potentially causative through the linkage analysis on the trio families. The pathway enrichment analysis was also provided to assess the potential interaction between genes identified in this study and previously reported together. The 5 novel identified overlap genes by both above mentioned test with the rare mutations account for an overall 20% (26 /133 patients) incidence, together with the 2 known genes together would account for an overall 20% incidence for non-obstructive azoospermia in this study. The study is limited by the lack of functional biological study.

**Conclusions:** Five novel genes were identified associated with non-obstructive azoospermia by means of both rare variant association study and linkage analysis through trio families. They could account for about 20% clinical incidence among the patients in our study.

**Patient summary:** 133 infertile patients (11 of them with parents enrolled) with idiopathic non-obstructive azoospermia and 300 fertile male controls were recruited from single clinic center.

All patients underwent semen analyses at least on three different occasions.

## 1. Introduction

Approximately 7% of male population worldwide suffer from infertility,^1^ which is mainly caused by azoospermia, a condition with the highest frequency of known genetic factors contributing to male infertility (about 25%).^2^ Given that nearly 50% of infertility cases are associated with genetic defects,^3^ the genetic etiology for azoospermia is still very elusive, the genetic basis of azoospermia in most cases remains unknown. In terms of pathobiological mechanism, azoospermia can be categorized into two groups: non-obstructive azoospermia which is defined as no sperm in the ejaculate due to failure of spermatogenesis, and obstructive azoospermia which is defined as the absence of spermatozoa in the ejaculate despite of the normal spermatogenesis.

Non-obstructive azoospermia (NOA) is the most severe form of azoospermia and lacking of effective cures, whereas patients with obstructive azoospermia usually can be treated with assisted reproductive technologies (ART) surgery and bypass infertility in this way. To date, mutations on several genes have been identified causative for, or associated with NOA, the acknowledged genes include X chromosome linked genes TEX11,^4–7^ FAM47C,^8^ MAGEB4,^9^ and AR;^10,11^ and autosomal genes PIWIL1,^12–15^ BRCA2,^16^ BOLL and DAZL,^17,18^ TEX15,^19–21^ SYCP3,^22^ NR5A1,^23^ SOHLH1,^24^ WT1,^25,26^ TAF4B and ZMYND15,^27^ FANCM,^28^ SPINK2,^29^ TEX14,^30^ DNAH6,^30^ MEIOB,^31^ RNF212 and STAG3,^32^ XRCC2,^33,34^ TDRD7,^35^ TDRD9,^36^ DMC1,^37^ SYCE1,^38^ NPAS2,^39,40^ MCM8,^41^ MEI1,^42,43^ and STX2.^44^ A surprisingly large proportion of total genes are specifically expressed in the male testis,^45^ suggesting that thousands of genes may involve in the spermatogenic process. Therefore, the physiological pathway complexity may contribute to the large number of unresolved idiopathic NOA patients.

However, currently tools for understanding the genetic etiology of NOA is very limited due to the lack of large pedigrees resulting from the infertility. In this study we performed whole exome sequencing (WES), a powerful tool for identifying the pathogenic factors for inherent diseases,^46,47^ on 133 unrelated patients and 300 independent fertile male controls for rare variant association study (RVAS), a kind of modified genome wide association study (GWAS);^48–55^ meanwhile 11 trio families (the proband with his parents) were also analyzed in a maternal inherited or a *de novo* pattern to identify the causative genes. Meanwhile proteins interaction pathway enrichment analysis was also performed to provide potential evidence to further address the abovementioned question.

## 2. Materials and methods

### 2.1. Study design and patients

This study was approved by the Medical Ethics Review Board of The First Hospital affiliated to Jilin University (Changchun, China), and with written informed consent from the patients and their parents, as well as the controls. 133 infertile patients (11 of them with their healthy parents enrolled together as trios) with idiopathic NOA (no sperm in the ejaculate even after centrifugation) and 300 fertile male controls were recruited from the Reproductive Medicine & Prenatal Diagnosis Center of The First Hospital, Jilin University (Changchun, China). All patients underwent semen analysis at least on three different occasions, and were excluded if be with a history of orchitis, obstruction of vas deferens or endocrine disorders. All the men from control group were fertile and had fathered at least one child.

We selected those affected by idiopathic NOA; 133 subjects and 11 trios signed the informed consent for this study and were enrolled. These patients were selected according to the following criteria: (i) azoospermia due to either spermatogenic arrest or Sertoli cell-only syndrome (SCOS; type I or II); (ii) normal karyotype; (iii) without absence of Y-chromosome microdeletions and TEX11 mutations (including the copy number variation (CNV) of TEX11); and (iv) absence of a list (Supplementary Table 1) of known causes of azoospermia. Clinical characteristics of the patients are reported in Table I.

### 2.2. Genomic analysis

The genomic DNA was prepared from the peripheral blood samples. All patients and the trio families enrolled in the study underwent WES sequencing performed by BGI Genomics (Shenzhen, China) according to previous report.^56^ Variants were annotated using ANNOVAR^57^ as loss of function (LOF) or missense variants. LOFs were defined as frameshift insertions or deletions, gain or loss of stop codon, insertions, deletions, and disruption of canonical splice site dinucleotides. CNVs were analyzed with ExomeDepth and CNVkit. Pathway enrichment analysis were performed with Cytoscape_v3.7.2 software based on the String database (https://string-db.org/). The data deposited in the CNSA (https://db.cngb.org/cnsa/) of CNGBdb with accession number CNP0001080.

### 2.3. Statistical analysis

Only LOFs with minor allele frequencies (MAF) ≤0.1% as per the Genome Aggregation Database (gnomAD) were considered for inclusion in the gene burden association testing, which was performed with SKAT. The p values reported are two-tailed. When applicable, multiple testing correction was performed using the FDR method and FDR < 0.05 was considered significant. Quantile-quantile plot (QQ plot) was achieved with the RVAS data and draw in R program. All analyses were performed using R v3.6.1 (www.R-project.org) and Bioconductor v3.4.

## 3. Results

The test flow for this study is shown in Figure 1, a total of 1021 NOA patients were enrolled in, of which 299 cases with abnormal karyotyping results were excluded, followed by 281 cases exclusion due to the AZF region microdeletion; after this, 305 cases were reluctant to accept further test and were excluded in the following study; then a list (Supplementary Table 1) of known genes of azoospermia were developed as custom genes panel screening test, 3 cases with positive results (2 with TEX11 mutation, 1 with AR mutation) were excluded in this step; eventually, 133 NOA patients without abnormal karyotyping, AZF region microdeletion, and the known causative genetic mutation were selected to perform WES; meanwhile 300 independent fertile men were selected as the control population in this study; among the 133 NOA cases, 11 were enrolled with their parents together as the trio family, linkage analysis for these 11 Trios were also performed to screen the causative genes.

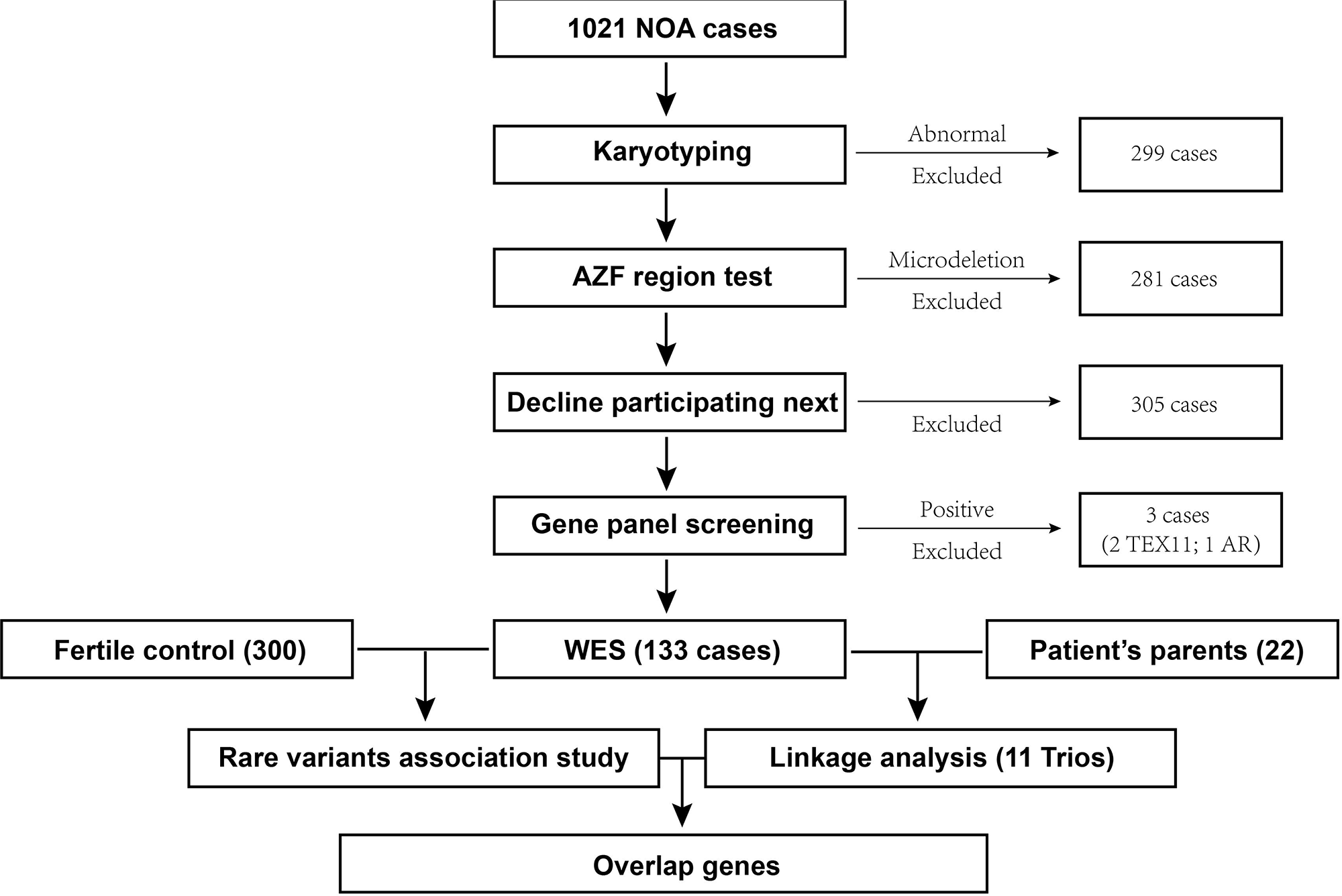

RVAS were analyzed for these cases and controls, the testis expressed genes with p value lower than 0.05 were labelled in color blue; meanwhile the genes in blue color were changed into yellow color if the genes were reported previously associated with NOA trait, or changed into red if the genes were overlapped identified in the linkage analysis for trios family. The quartile-quartile plot for the RVAS analysis was also shown in Figure 2. Two previously reported genes, FAM47C and BRAC2 (Table 2), were identified associated with NOA in this study; and five novel genes were revealed in both the RVAS (Table S2) and trios linkage analysis (Table 3) study.

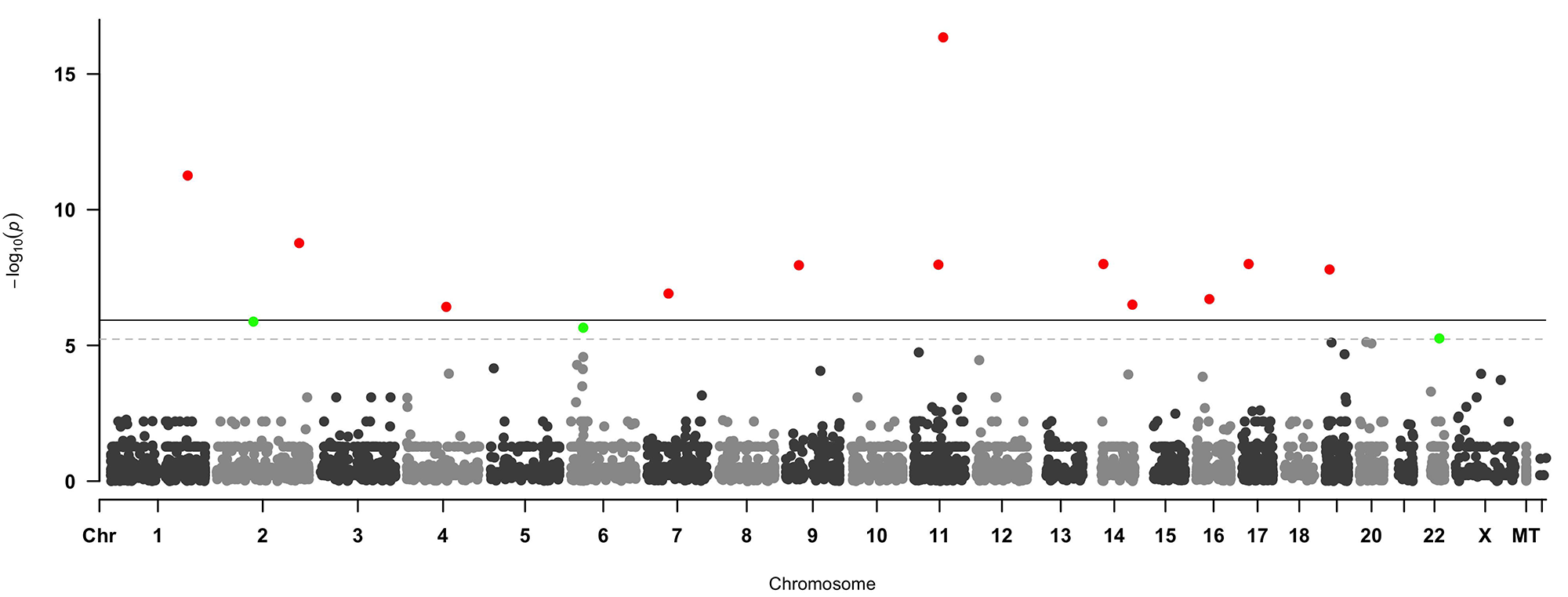

The RVAS identified associated genes, as well as the previously reported genes, were analyzed for the pathway enrichment in Figure 3. Previously reported genes were labelled in green color, interestingly, previously reported genes associated with NOA, except MAGEB4, FAM47C, SPINK2 and STX2, tend to accumulate in the meiosis related protein interaction pathway which is centered around PIWIL1, TEX11, TEX15, DAZL, TEX14, etc. A part of the RVAS identified genes also involved in the enriched pathway and provided functional potentials to be associated with NOA; meanwhile a small part of genes, like HLA-A, tend to be enriched in the immunity pathway, however no previous reports or linkage analysis evidence can support the association between the immunity pathway we identified through RVAS analysis and NOA trait. Since FAM47C was identified as highly associated with NOA previously, the FAM47C mutation (frameshift mutation similar with previous report) from patient 58 in our study was also identified as the genetic etiology accordingly.

Trios family WES data were analyzed and summarized in Table 3 according to a maternal inherited pattern or the *de novo* mutation mechanism, paternal inherited variations were excluded, also the probands’ variations were preserved in the summary table 3 only if the variations were not found in the 300 male control population. All of the WES identified variations were validated with sanger sequencing in this Table 3.

## 4. Discussion

Combining the data of RVAS analysis and the trio family analysis show the overlapped genes may associated with NOA. Especially, genes also involved in the enriched pathway with previously reported genes, suggesting a high potential to be associated with and causative for NOA. The previous reported genes, FAM47C and BRCA2, were also identified in this study, the FAM47C mutation (frameshift mutation similar with previous report) from patient 58 and the BRCA2 mutation (insertion similar with previous report) from patient 102 in our study was also identified as the genetic etiology accordingly. Nevertheless, the limitation of this study lies in that prospectively more evidence like the functional cellular or physiological experiments could be needed to address this question better, and that the missense mutation which may be pathogenic was not taken into account in this study.

The variations can be classified as Likely Pathogenic according to ACMG guidelines,^58^ PS2 (strong evidence) and PS4 (strong evidence) and PM2 (moderate evidence) and PM4 (moderate evidence).

## 5. Conclusion

We demonstrate the association study between the 133 NOA patients and 300 fertile men control, as well trio families were also analyzed to further investigate the causative genes for NOA trait. We conclude that the overlapped identified genes and may associated with NOA. The other RVAS or trio analysis identified genes, especially the genes involved in the enriched pathway may also associated with NOA. This study provides a potential possibility for better diagnosis in the future clinical application.

## Data Availability

The data deposited in the CNSA (https://db.cngb.org/cnsa/) of CNGBdb with accession number CNP0001080.

